# Cruise Control Study: Simplification of IVF Monitoring in a Mixed Protocol Using a Novel Dosing Regimen

**DOI:** 10.1101/2023.10.23.23297336

**Authors:** Jonas Benguigui, Eva Kadoch, Amro Bannan, Simon Phillips, Robert Hemmings, François Bissonnette, Isaac-Jacques Kadoch

## Abstract

**Objective:** To identify the subset of the in vitro fertilization (IVF) population suitable for minimal monitoring by implementing a novel dosing regimen.

**Methods:** A retrospective study conducted between April 2021 and August 2022. Eligible participants were aged 18 or older, had undergone IVF stimulation using an antagonist protocol, and were prescribed a combination of follitropin delta and human menopausal gonadotropin. The dosage was either based on a patient-specific dosing regimen developed by the ovo clinic utilizing weight and AMH levels (Group 1, n=356) or determined through clinical evaluation by the physician (Group 2, n=358). On day 6, ultrasound and serum hormone analyses were performed, with adjustments made solely to the menotropin dosage in necessary.

**Results:** The study enrolled a total of 714 patients. In Group 1, 80,3% of patients were stimulated at maximal doses compared to 14,5% in Group 2. No cases of moderate or severe cases of ovarian hyperstimulation syndrome (OHSS) were recorded. The frequency of dose adjustments before day 10 was minimal. Patients treated with non-maximal doses according to the dosing regimen showed significantly fewer adjustments on day 6 compared to those treated according to physician’s assessment (24.6% versus 46.9%, p<0.001). Among this subgroup, OHSS risk was observed in 30.4% of cases.

**Conclusion:** Our innovative dosing regimen suggests that initial monitoring on day 10 would suffice for IVF patients with low ovarian reserve undergoing maximal stimulation.

## INTRODUCTION

In the context of ovarian stimulation for in vitro fertilization (IVF), studies have highlighted the effectiveness and safety of personalizing the prescription of follitropin delta based on individual patient factors such as weight and anti-Müllerian hormone (AMH) levels (Nyboe Andersen *et al*., 2017). This personalized approach has proven to be superior to conventional ovarian stimulation methods, leading to a reduction in the risks associated with failed stimulation and ovarian hyperstimulation syndrome (OHSS) during IVF cycles (Nyboe Andersen 2017).

The interplay between follicle-stimulating hormone (FSH) and luteinizing hormone (LH) bioactivity has been extensively studied and studies have demonstrated that the combined use of follitropin delta and highly purified human gonadotropins (HP-hMG) yields a higher number of retrieved oocytes and a higher rate of blastulation compared to the administration of follitropin delta alone (Bissonnette *et al*., 2021). Given the robust evidence, our center has readily implemented and incorporated this combination protocol, wherein the dosing algorithm for follitropin delta is harmonized with a calculated equivalent dose of HP-hMG, a dosage determination established by the ovo clinic.

From the patient’s standpoint, undergoing IVF is a physically and emotionally taxing journey. The process involves numerous examinations that not only burden patients but can also pose as barriers to accessing the treatment. Additionally, the need for repeated ultrasounds can lead to additional financial burdens for patients.

With these considerations in mind, the objective of our study was to identify the eligible patient population that can benefit from a streamlined IVF dosing regimen. This regimen aims to decrease the frequency of ultrasounds and blood tests, specifically by eliminating the need for a day 6 visit, all while ensuring that the quality of treatment outcomes remain uncompromised.

## MATERIAL AND METHODS

### Patient Selection

This retrospective study was conducted at the ovo clinic in Montreal, Canada, from April 2021 to August 2022. All patients aged 18 years or older undergoing IVF stimulation cycles using an antagonist protocol and receiving a mixed protocol of follitropin delta and HP-hMG were included in the study. Exclusion criteria included patients with a single ovary, those undergoing a second stimulation cycle with the DuoStim protocol (Massin *et al*., 2023), and those undergoing oocyte preservation cycles.

### Choice of Gonadotropin Doses Based on the Dosing Regimen

Regarding the prescription of follitropin delta, an algorithm based on the patient’s weight and recent AMH levels was used. The maximum daily dose of follitropin delta (Rekovelle®; Ferring Pharmaceuticals) was 12 micrograms for women with an AMH level below 2.1 ng/L, regardless of their weight. For women with an AMH ≥ 2.1 ng/L, the daily dose decreased from 0.19 to 0.10 micrograms/kg according to the AMH concentration (Table 1). The dose was rounded to the nearest 0.33 micrograms to correspond to the dosage scale on the injection pen.

**Table 1:**
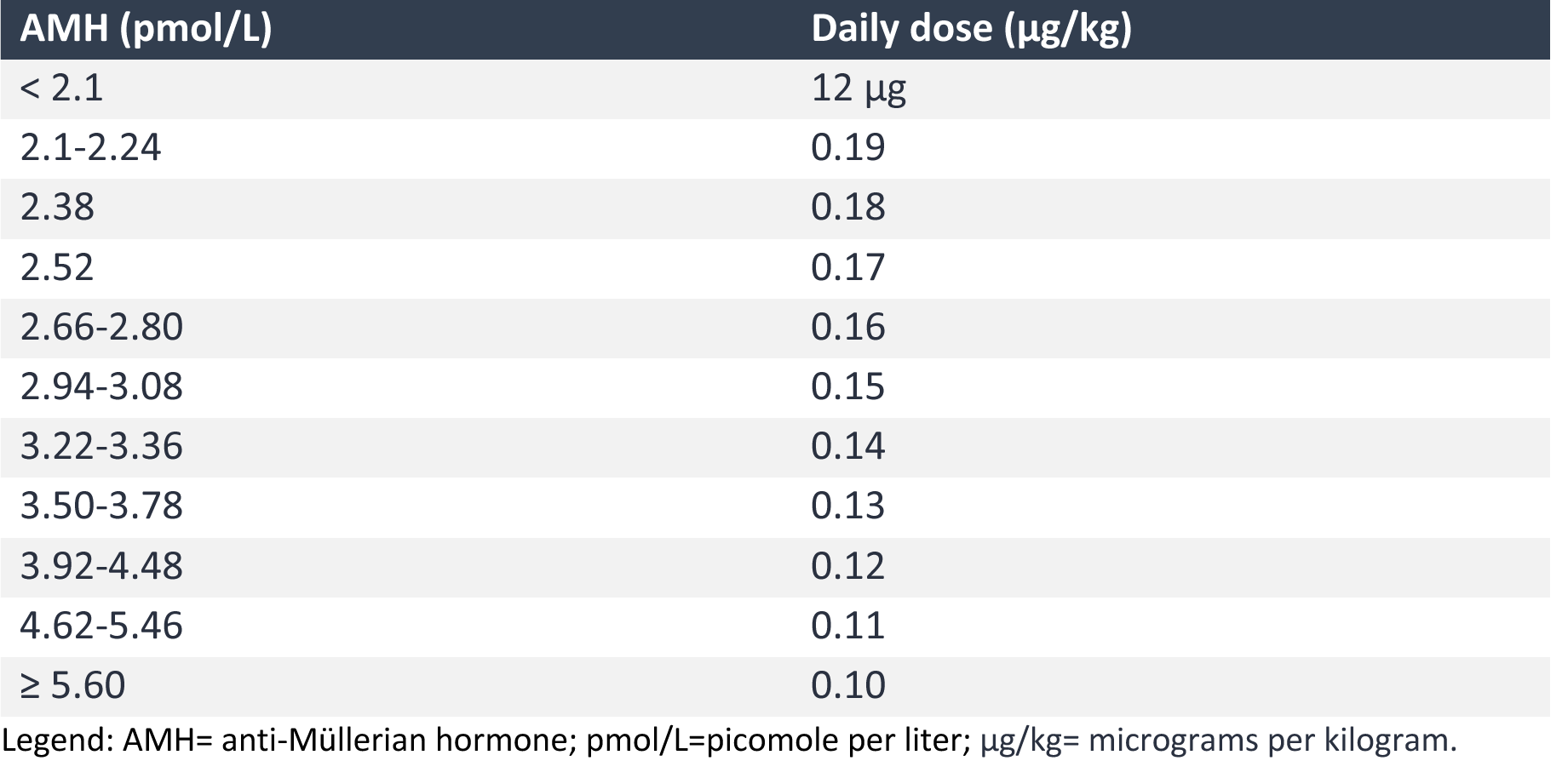
Follitropin delta dosing algorithm.

| AMH (pmol/L) | Daily dose (µg/kg) |
| --- | --- |
| < 2.1 | 12 µg |
| 2.1-2.24 | 0.19 |
| 2.38 | 0.18 |
| 2.52 | 0.17 |
| 2.66-2.80 | 0.16 |
| 2.94-3.08 | 0.15 |
| 3.22-3.36 | 0.14 |
| 3.50-3.78 | 0.13 |
| 3.92-4.48 | 0.12 |
| 4.62-5.46 | 0.11 |
| ≥ 5.60 | 0.10 |

Legend: AMH= anti-Müllerian hormone; pmol/L=picomole per liter; µg/kg= micrograms per kilogram.

In our clinic, some physicians adhered strictly to the prescription using the dosing regimen (“Dosing Regimen” group) while other physicians tended to prescribe the mixed protocol using their clinical assessment (“Clinical Assessment” group). For HP-hMG (Menopur®; Ferring Pharmaceuticals), the dose was established according to an equivalence that we defined internally based on the follitropin delta dosing algorithm (Table 2).

**Table 2.** Choice of HP-hMG Dose According to Follitropin Delta Dosing Algorithm.

| Follitropin delta dose (µg) | Choice of HP-HMG dose (IU) |
| --- | --- |
| Up to 5.66 | 75 |
| Between 6 and 8.66 | 150 |
| Between 9 and 11.66 | 225 |
| 12 | 300 |

Legend: HP-hMG=Highly purified human menopausal gonadotropin; IU=International Units; µg= micrograms.

### Stimulation Protocol

Prior to ovarian stimulation, all patients underwent a systematic preparatory phase involving estradiol administration (17 beta-estradiol 2 mg) over a span of 7 to 10 days. This preparation aimed to establish a reduction of the follicular size discrepancy and to allow a better cycle scheduling (Fanchin et al., 2003). Following this, the administration of gonadotropins for ovarian stimulation could begin.

On the sixth day of stimulation, a preliminary ultrasound assessment and estradiol level measurement were conducted. Ovarian response was considered optimal when the estradiol level was between 2000 and 4000 pmol/L. If the estradiol level was below 2000 pmol/L, an increase in HP-hMG dose could be considered, while a decrease in dose could be performed if the estradiol level was above 4000 pmol/L. Moreover, the introduction of a gonadotropin-releasing hormone (GnRH) antagonist was variable, contingent upon the attainment of a follicle size of 14 mm or an estradiol level surpassing 2000 pmol/L. The antagonist regimen persisted until the point of ovulation triggering. Subsequent monitoring was contingent upon the follicle size observed on day 6. The standard criteria for ovulation triggering included the presence of at least 3 pre-ovulatory follicles, each measuring between 16 and 22 millimeters in diameter.

Ovulation was triggered either through 5000 units of hCG (Ferring Pharmaceuticals) or 1 mg of Busereline (Suprefact®; Xediton Pharmaceuticals), the latter applied to cases involving embryo freeze-all. In select cases, a double triggering method was used involving both 5000 IU hCG and Busereline 1 mg. Oocytes were collected 36 hours post-triggering via transvaginal ultrasound-guided follicular aspiration.

### Statistical Analysis

Continuous variables were compared between independent groups after evaluating normal distribution using the T test, ANOVA, and linear regression. For dichotomous variables, the Chi-square test and linear regression were used. A p-value less than 0.05 resulted in rejection of the null hypothesis and significant difference between the groups. Statistical analyses were performed using IBM SPSS, version 26.0.

### Ethical Approval

The study was approved by Veritas IRB, an independent research ethics committee. Study tracking number: 2023-3189-13659-2.

## RESULTS

### Population Distribution

A total of 772 cycles were included. The population selection is summarized in the flow diagram in Figure 1. The percentage of cancelled cycles was 7.5% (n=58), 81.0% of which were cancelled due to inadequate response (n=47) and 19.0% (n=11) due to other reasons. Among the hypo-response cases, the mean age of patients was 38.7±3.1 years and the mean AMH level was 0.81±1 ng/mL.

**Figure 1.**
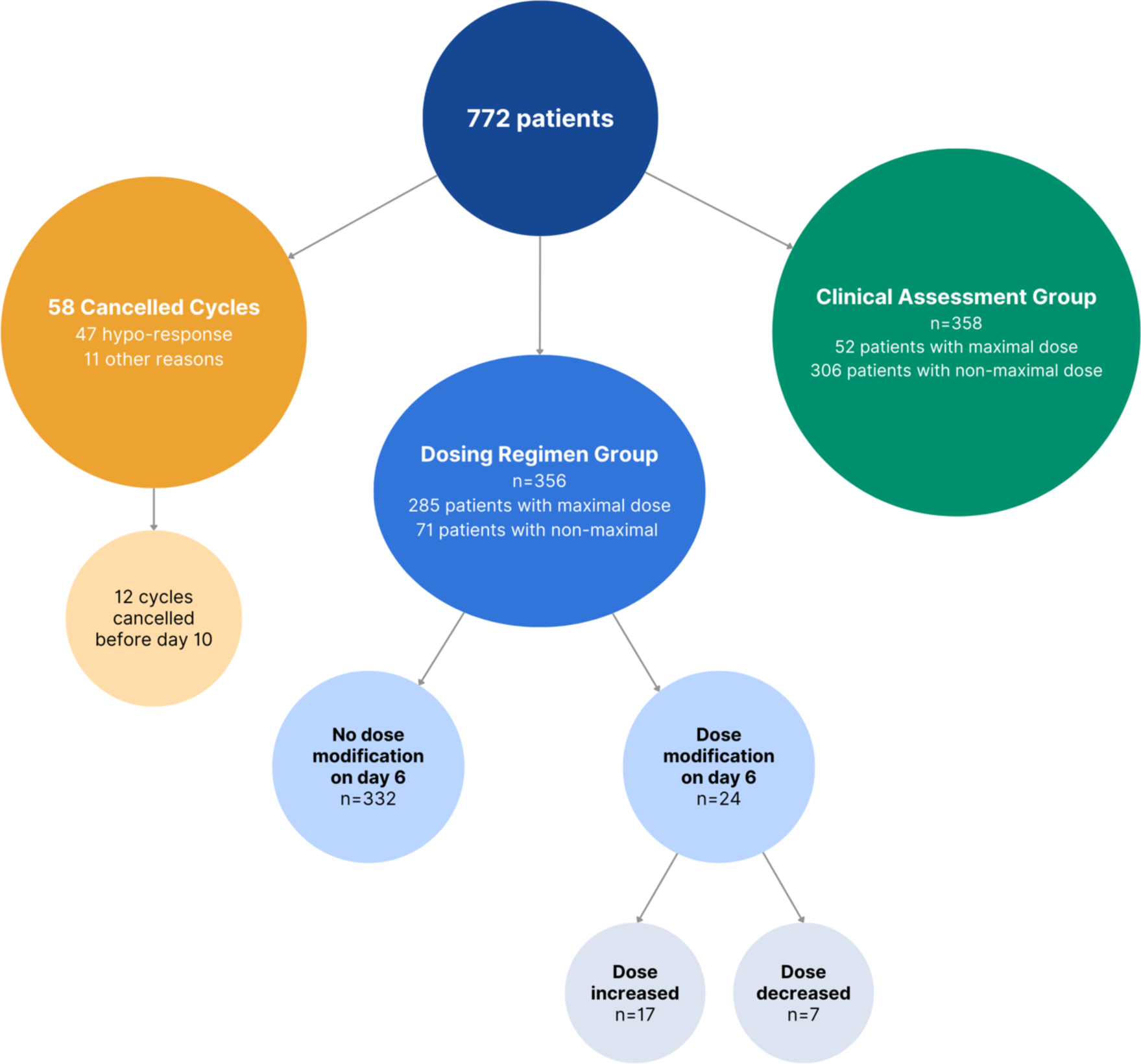
Flow diagram.

### Comparison of Dosing Regimen and Clinical Assessment Populations

In the “Dosing Regimen” group (n=356), 93.2% of patients maintained the same dose until the point of ovulation trigger. Notably, only 24 patients experienced a dose modification, with 17 patients had an increase in dosage, while 7 patients saw a reduction. A detailed synopsis of the demographics between Group 1 and Group 2 is provided in Table 3 for reference.

**Table 3.** Comparison of population characteristics with gonadotropin prescription according to the dosing regimen (group 1) or not (group 2).

|  | Group 1<br>(n=356) | Group 2<br>(n=358) | p-value |
| --- | --- | --- | --- |
| Age, years (mean ± SD) | 37.4±4.2 | 34.9±4.6 | <0.05 |
| Rank of attempt (mean ± SD) | 1.7±1.5 | 1.4±1.1 | <0.05 |
| AMH, ng/mL (mean ± SD) | 1.98±2.6 | 3.4 ±2.4 | <0.05 |
| Weight, kg, (mean ± SD) | 68.4±16.9 | 67.5±16.5 | NS |
| Maximum dose used (%) | 80.3 | 14.5 | <0.05 |
| HP-hMG doses, IU, (mean ± SD) | 234±75 | 182±74 | <0.05 |
| Follitropin delta doses, IU, (mean ± SD) | 10.5±2.9 | 10.3±1.8 | NS |
| Monitoring per patient (mean ± SD) | 2.8 ±0.7 | 2.6±0.6 | <0.05 |
| Dose modification on day 6 (%) | 5.6 | 41.6 | <0.05 |
| Dose increase on day 6 (%) | 4.2 | 36 | <0.05 |
| Dose decrease on day 6 (%) | 1.4 | 5.6 | <0.05 |
| Estradiol level on day 6, pmol/L, (mean ± SD) | 1186±1006 | 1448±1175 | <0.05 |
| Trigger day, (mean ± SD) | 12±1.6 | 12±1.2 | NS |
| Triggers before day 10, % (n) | 4.5% (16) | 3.1% (11) | NS |
| Mature oocytes retrieved (mean ± SD) | 10.7±7.5 | 15.1±8.6 | <0.05 |
| Blastocysts (mean ± SD) | 4.6±3.7 | 6.3±4.7 | <0.05 |

Legend: SD=Standard Deviation; AMH=anti-Müllerian hormone; kg=kilogram; ng/mL=nanogram per milliliter; HP-hMG=highly purified human menopausal gonadotropin; NS=Not significant.

Regarding Group 1, a few notable distinctions emerged. This cohort exhibited an advanced age (37.4±4.2 vs. 34.9±4.6 years) in comparison to Group 2. Additionally, Group 1 displayed lower AMH levels (1.98±2.6 vs. 3.37±2.4 ng/mL), coupled with higher average doses of HP-hMG (234±75 vs. 182±74 IU). Notably, a disparity in the yield of mature oocytes and blastocysts was evident, with Group 1 recording lower counts (10.7±7.5 vs. 15.1±8.6 and 4.6±3.7 vs. 6.3±4.7, respectively). In terms of patient weight, average doses of follitropin delta, and stimulation duration, no significant difference surfaced between the two groups. The timing of ovulation trigger remained consistent around day 12 for both groups.

Within Group 1, there was a notable overrepresentation of individuals receiving the maximum gonadotropin dosage (HP-hMG 300 IU and follitropin delta 12 μg) compared to Group 2 (80% vs. 14.5%). In this population, 3.8% (n=13) opted for embryo freeze-all due to the risk of OHSS. However, no moderate or severe OHSS incidents were observed in this subgroup.

### Ovulation Monitoring and Trigger in the Low Ovarian Reserve Population

In the population receiving maximal dosage, a total of 940 blood tests and ultrasound monitorings were performed during ovarian stimulation, including 439 prior to day 10. In this group, the gonadotropin doses were modified only on day 6 of stimulation in 1.7% of cycles. The percentage of patients in this population who underwent ovulation triggering before day 10 was 3.3%.

### Non-Maximum Dose Population

Due to the overrepresentation of the maximum dose population in Group 1 (80.3%) and given that this group exhibited minimal need for dose adjustments, the decision was made to create two additional groups by excluding patients receiving the maximum doses. These new groups were designated as Group A (“Dosing Regimen” group) and Group B (“Clinical Assessment” group) (Table 4).

**Table 4.** Characteristics of cycles according to the dosing regimen prescription or not in the non-maximal dose population.

|  | Group A:<br>Dosing<br>Regimen<br>(n=71) | Group B:<br>Clinical<br>Assessment<br>(n=307) | p-value |
| --- | --- | --- | --- |
| Age, years (mean ± SD) | 34.4±4.5 | 34.2±4.4 | NS |
| AMH, ng/mL (mean ± SD) | 4.8±4.6 | 3.4±2.6 | **<0.05** |
| Weight, kg, (mean ± SD) | 62.8±15.4 | 70±15.7 | **<0.05** |
| HP-hMG doses, IU, (mean ± SD) | 166±47 | 160.5±61 | 0.48 |
| Follitropin delta doses, IU, (mean ± SD) | 8.4±1.8 | 10.0±2.2 | **<0.05** |
| HP-hMG dose suggested by the dosing regimen (mean ± SD) | 166±47 | 208±66.5 | |
| Follitropin delta doses suggested by algorithm (mean ± SD) | | 9.3±2.2 | |
| Monitorings per patient (mean ± SD) | 2.5±0.67 | 2.67±0.67 | NS |
| Dose modification on day 6 (%) | 26.7 | 55.7 | **<0.05** |
| Dose increase on day 6 (%) | 18.3 | 49.5 | **<0.05** |
| Dose decrease on day 6 (%) | 8.4 | 5.9 | NS |
| Trigger day | 11.1±1.5 | 11.9±1.4 | NS |
| Freeze all for a risk of OHSS (%) | 43.7 | 27.4 | **<0.05** |
| Mature oocytes retrieved (mean ± SD) | 15.6±6.9 | 12.3±6.9 | **<0.05** |
| Blastocysts (mean ± SD) | 8.4±4.4 | 6.6±4.4 | **<0.05** |

Legend: $\mu\pm$ SD: mean ± standard deviation; AMH: anti-Müllerian hormone; HP-hMG: highly purified human menopausal gonadotropin. OHSS: Ovarian hyperstimulation syndrom; NS: Non-Significant.

Group B had prescribed doses of HP-hMG that were significantly lower than those suggested by the established dosing regimen (160.5±61 vs. 208±66.5 IU; p<0.01), while the prescribed follitropin delta doses were comparable to those suggested by the algorithm (10.0±2.2 vs. 9.3±2.2 μg).

Comparatively, Group A had significantly higher AMH levels (4.8±4.6 vs. 3.4±2.6 ng/mL) along with a greater number of retrieved mature oocytes and obtained blastocysts (15.6±6.9 vs. 12.3±6.9 and 8.4±4.4 vs. 6.6±4.4, respectively). Dose modifications were significantly fewer in Group A (26.7% vs. 55.7%).

Interestingly, the risk of OHSS was significantly higher in the non-maximal dose population as compared to the maximal dose group (30.4% vs. 4.7%).

Figure 2 represents the distribution of estradiol levels on day 6 across the two groups. Group A demonstrated a more frequent achievement of the target estradiol level on day 6 (39.4% vs 16.9%; p<0,05). Conversely, in Group B, a larger proportion of patients had estradiol levels below 2000 pmol/L on day 6 (80.3% vs 53.5%), resulting in more frequent increases in gonadotropin doses (29.7%). Notably, gonadotropin doses were reduced in 7.4% of cycles with estradiol levels between 2000 and 4000 pmol/L and in 50% of cycles with levels exceeding 4000 pmol/L. In group A, a total of 45.7% of patients (n=32) underwent freeze-all for OHSS risk.

**Figure 2.**
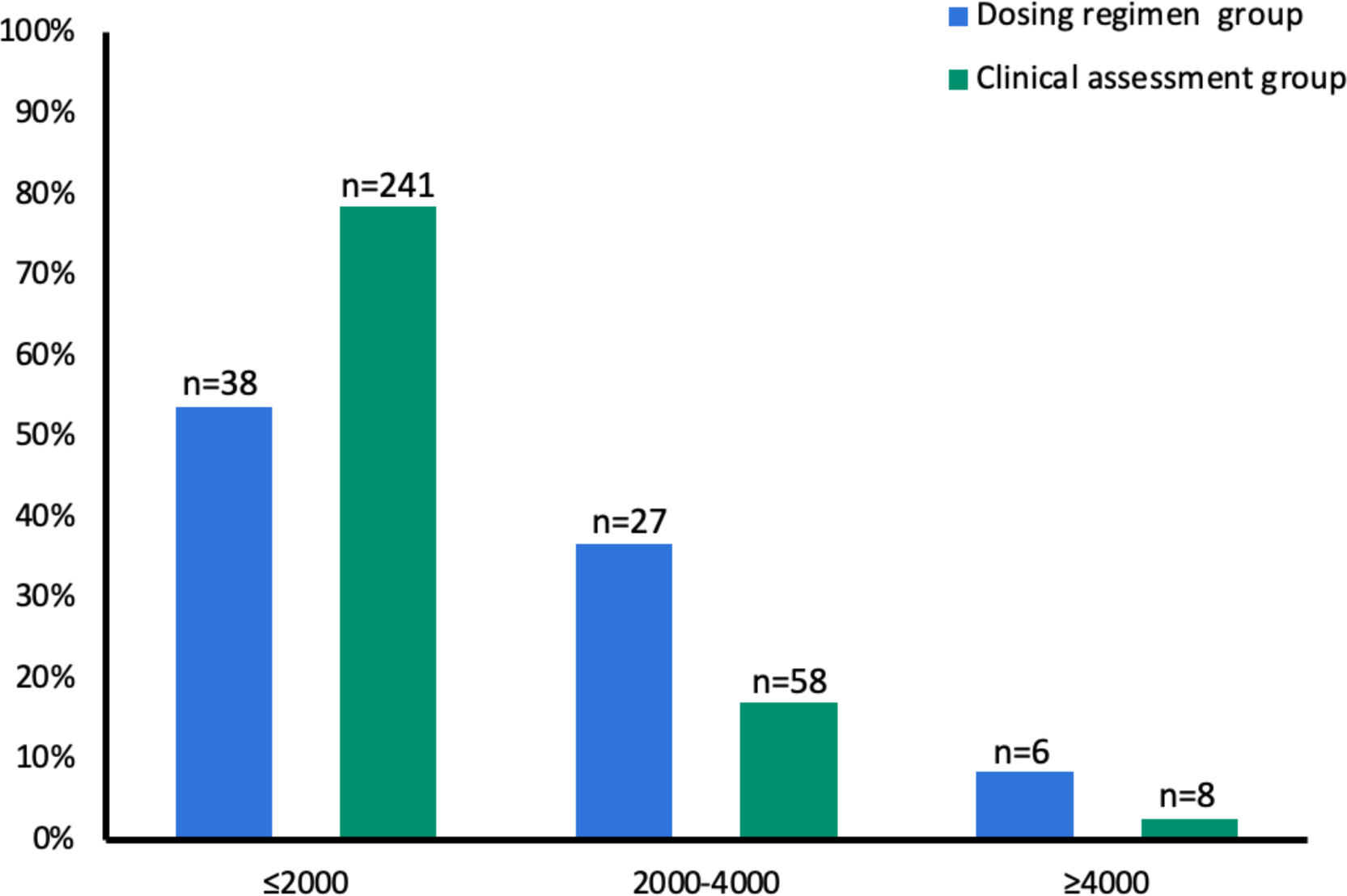
Distribution of the population according to estradiol levels (pmol/L) on day 6 of ovarian stimulation in the non-maximal dose population in the dosing regimen group (A) (n=71) and clinical assessment group (B) (n=307).

## DISCUSSION

This study proposes a prospective shift in the approach to monitoring IVF treatment for patients with low ovarian reserve. Experiencing infertility and undergoing IVF can be emotionally taxing, and the numerous clinic visits for tests add to the stress and expenses for couples. While these tests are essential for mitigating the risks of unsuccessful IVF and OHSS, our objective was to explore the feasibility of simplifying this process.

Our previous research indicated that individualized dosage in a mixed protocol of follitropin delta and HP-hMG based on individual needs could lead to better outcomes in IVF. This approach led to an increased number of viable blastocysts on days 5 and 6 as well as a significantly reduced OHSS incidence (Bissonnette *et al*., 2021). This suggests that personalized gonadotropin dosing can effectively limit the risks of OHSS while ensuring the effectiveness of the protocol. This led us to re-evaluate the necessity of frequent monitoring, considering the burden and challenges associated with these exams. A Cochrane review (Kwan *et al*., 2021) demonstrated that combined ultrasound and hormonal tests were not more effective than ultrasound monitoring alone as evidenced by enhanced clinical pregnancy rates and OHSS incidence. However, the review included studies of moderate quality, characterized by limited evidence. Unlike our study, this review did not segment patients based on ovarian reserve and stimulation protocols.

In our study, the group that followed the prescribed dosing regimen consisted of older individuals with notably lower AMH levels. As anticipated, this group received higher gonadotropin dosages, resulting in a lower number of retrieved oocytes and blastocysts. Moreover, dose adjustments on day 6 were significantly lower within this cohort, potentially attributed to certain physicians’ infrequent use of the proposed dosing regimen. In case of low ovarian reserve, maximal doses of HP-hMG and follitropin delta, equivalent to the doses recommended by the algorithm, were prescribed. Prior research has indicated that gonadotropin prescriptions surpassing 150 units per day (van Tilborg *et al*., 2017) or 300 units per day (Siristatidis and Hamilton, 2007) do not improve pregnancy probabilities compared to higher doses. Consequently, the increase in doses on day 6 for patients already receiving maximal dosage might not significantly affect the overall treatment plan.

Furthermore, the risk of OHSS was minimal within this specific population (4.7 %). Notably, diagnosing OHSS within contexts of low ovarian reserve can be controversial. Given the very low rate of dose adjustments on day 6 and the minimal OHSS risk, it is reasonable to question the necessity of monitoring prior to day 10 in this population. Our results indicated that 439 ultrasounds and blood tests, representing 46.7% of all monitoring sessions in the general population, could have been averted.

During the study period, a 7.5% cycle cancellation rate was observed, predominantly among patients with low ovarian reserve receiving maximal doses. Among the 47 cycles cancelled due to hypo-response, only 12 were cancelled on days 6 or 7, suggesting a tendency among physicians to extend treatment for a few days before considering cancellation. However, early day 10 monitoring could potentially lead to needless treatment prolongation by 3 to 4 days.

The study raises questions about the relevance of early monitoring for introducing a GnRH antagonist in patients with diminished ovarian reserve. However, a recent randomized controlled study (Luo *et al*., 2021) showed that the early and fixed introduction of this antagonist had no effect on retrieved oocytes numbers compared to a variable introduction. This analysis excluded patients with polycystic ovary syndrome (PCOS) and confirmed the results of a previous meta-analysis (Al-Inany *et al*., 2005), which also revealed that fixed or variable antagonist introduction did not significantly influence pregnancy rates or premature ovulation rates.

Given the overrepresentation of the diminished ovarian reserve population and thus the prevalence of maximal dosage, this subgroup was excluded from further analysis. In the “Dosing Regimen” group (n=71), AMH levels were significantly higher. This could be attributed to physicians’ tendency to use the prescription dosing regimen more often in cases of higher ovarian reserve, with the goal of adjusting gonadotropin levels to reduce the risk of OHSS. Nonetheless, these findings warrant cautious interpretation due to the relatively limited patient count.

In our clinic, our standard practice involves adjusting gonadotropin doses on day 6 based on each patient’s ultrasound and blood test responses. Estradiol levels below 2000 pmol/L signify suboptimal response and the doses are then increased at the physician’s discretion. Conversely, estradiol levels above 4000 pmol/L signal hyper-response and potential OHSS risk, necessitating dose reduction. Our study revealed that estradiol levels below 2000 pmol/L were more prevalent in the “Clinical Assessment” group (80.3%) compared to the “Dosing Regimen” group (53.5%). In fact, in Group B, HP-hMG doses prescribed were lower than the dosing regimen’s suggestions (160.5±61 vs. 208±66.5 IU), possibly reflecting physician’s inclination to initiate treatment with lower gonadotropin doses to avert potential OHSS risk. Applying the dosing regimen could enhance oocyte and blastocyst yields without increasing OHSS risk.

Within the population receiving non-maximal doses, dose adjustments on day 6 were noted at 26.7% and 55.7% in Groups A and B, respectively. Furthermore, OHSS risk was more prevalent in this subgroup. These results emphasize the importance of day 6 monitoring for this population. Relying solely on blood tests on day 6, with gonadotropin dose adjustments based on estradiol levels, seems reasonable. For estradiol levels higher than 4000 pmol/L, ultrasound monitoring on the next day could confirm the absence of mature follicles and trigger readiness.

Efforts have been made to streamline monitoring during IVF cycles. A recent study (Letterie *et al*., 2022) involving 1,591 IVF cycles explored leveraging artificial intelligence (AI) to optimize workflow by minimizing monitoring during ovarian stimulation in IVF. The AI algorithm, considering patient characteristics (age, FSH, estradiol, AMH, number of antral follicles, and body mass index), determined the ideal ovulation monitoring and triggering day for maximal mature oocyte retrieval. While the use of AI in fertility treatment is still relatively new, it holds potential to aid practitioners in managing IVF protocol monitoring, alleviating care burden.

To our knowledge, our study suggests for the first time that early monitoring before day 10 (coupled with standard initiation of antagonist) during ovarian stimulation might be avoidable for a substantial population of patients, without compromising IVF outcomes. This paradigm shift could profoundly alter fertility centers’ operations by reducing unnecessary examinations. This approach would significantly reduce patient burden, diminish work absenteeism, avoid redundant invasive tests during the protocol. Moreover, it would yield substantial financial benefits for both patients and medical professionals.

The study was conducted employing a prescription protocol based on an algorithm that has exhibited success across multiple IVF studies (Doroftei *et al*., 2023; Qiao *et al*., 2021). The combination of follitropin delta and HP-hMG has also been studied and proved to be a safe solution for patients, while increasing the number of collected oocytes and blastocysts obtained (Bissonnette *et al*., 2021). Standardizing gonadotropin prescriptions aimed to reduce bias related to subjective gonadotropin choices by physicians chosen more subjectively by the physician.

Nonetheless, contradictory results were observed compared to a recent study (Robertson *et al*., 2021). This study, conducted during the COVID-19 pandemic, aimed to optimize follicular monitoring strategies during IVF by minimizing checks. Results indicated that an ultrasound on day 5 of ovarian stimulation was crucial for predicting trigger day and hyper-response risk. However, this study was retrospective and did not stratify patients based on ovarian reserve, unlike our study which specifically targets patients with low ovarian reserve. Therefore, for patients not at risk of OHSS, bypassing early (day 6) monitoring is indeed feasible.

It is important to acknowledge the limitations of the study. The retrospective nature of the study inherently presents a potential bias to the findings. Additionally, the study population was primarily composed of older patients with diminished ovarian reserve. Although it represents a reflection of our general population in IVF, efforts were made to mitigate this bias through subgroup analysis excluding maximum-dose patients, but this resulted in small sample sizes with contrasting group characteristics. Furthermore, ongoing pregnancy were not intentionally assessed due to potential biases linked to preimplantation genetic testing for aneuploidy (PGT-A), endometrial preparation protocol variability, or patients with repeated implantation failure.

In conclusion, this study suggests that in cases of standardized prescription using a dosing regimen, early monitoring of ovulation before day 10 during ovarian stimulation can be omitted for women with diminished ovarian reserve with maximum doses of gonadotropins, all while upholding the efficacy of ovarian stimulation outcomes. This approach offers significant advantages to patients in terms of an enhanced IVF experience, encompassing improved time and cost efficiency, as streamlines practices for physicians. For women with normal or increased ovarian reserve, we propose initiating monitoring with a blood test on day 6, followed by an ultrasound on the subsequent day based on the results. However, further prospective studies are needed to validate these findings and assess the long-term impact on IVF outcomes.

## Attestation Statement

Data regarding any of the subjects in the study has not been previously published.

## Data Availability Statement

Data will be made available to the editors of the journal for review or query.

## Approvals

The study was approved by the Veritas Independent Review Board.

## Trial Registration

NCT05737979

## Date of Trial Registration

February 21^st^, 2023

## Declaration of Generative AI and AI-Assisted Technologies in the Writing Process

During the preparation of this work, the authors used ChatGPT in order to improve language and sparingly assist in the translation of text to English. After using this tool, the authors reviewed and edited the content as needed and take full responsibility for the content of the publication.

## ABBREVIATIONS

AI: Artificial Intelligence
AMH: Anti-Mullerian Hormone
ANOVA: Analysis of Variance
FSH: Follicle Stimulating Hormone
GnRH: Gonadotropin-Releasing Hormone
hCG: Human Chorionic Gonadotropin
HP-hMG: Highly Purified Human Gonadotropins
IU: International Unit
IVF: In Vitro Fertilization
LH: Luteinizing Hormone
PCOS: Polycystic Ovary Syndrome
PGT-A: Preimplantation Genetic Testing for Aneuploidy
OHSS: Ovarian Hyperstimulation Syndrome

## REFERENCES

1. Al-Inany H, Aboulghar MA, Mansour RT, Serour GI. Optimizing GnRH antagonist administration: meta-analysis of fixed versus flexible protocol. Reprod Biomed Online 2005;10:567–570.

2. Bissonnette F, Minano Masip J, Kadoch I-J, Librach C, Sampalis J, Yuzpe A. Individualized ovarian stimulation for in vitro fertilization: a multicenter, open label, exploratory study with a mixed protocol of follitropin delta and highly purified human menopausal gonadotropin. Fertil Steril 2021;115:991–1000.

3. Doroftei B, Ilie O-D, Anton N, Marcu O-A, Scripcariu I-S, Ilea C. A Narrative Review Discussing the Efficiency of Personalized Dosing Algorithm of Follitropin Delta for Ovarian Stimulation and the Reproductive and Clinical Outcomes. Diagnostics (Basel) 2023;13:177.

4. Fanchin, R., Salomon, L., Castelo-Branco, A., Olivennes, F., Frydman, N., & Frydman, R. (2003, Dec). Luteal estradiol pre-treatment coordinates follicular growth during controlled ovarian hyperstimulation with GnRH antagonists. Hum Reprod, 18(12), 2698–2703. 10.1093/humrep/deg516

5. Kwan I, Bhattacharya S, Woolner A. Monitoring of stimulated cycles in assisted reproduction (IVF and ICSI). Cochrane Database Syst Rev 2021;4:CD005289.

6. Letterie G, MacDonald A, Shi Z. An artificial intelligence platform to optimize workflow during ovarian stimulation and IVF: process improvement and outcome-based predictions. Reprod Biomed Online 2022;44:254–260.

7. Luo X, Pei L, Li F, Li C, Huang G, Ye H. Fixed versus flexible antagonist protocol in women with predicted high ovarian response except PCOS: a randomized controlled trial. BMC Pregnancy Childbirth 2021;21:348.

8. Massin N, Abdennebi I, Porcu-Buisson G, Chevalier N, Descat E, Piétin-Vialle C, Goro S, Brussieux M, Pinto M, Pasquier M, et al. The BISTIM study: a randomized controlled trial comparing dual ovarian stimulation (duostim) with two conventional ovarian stimulations in poor ovarian responders undergoing IVF. Hum Reprod 2023:dead038.

9. Nyboe Andersen A, Nelson SM, Fauser BCJM, García-Velasco JA, Klein BM, Arce J-C, ESTHER-1 study group. Individualized versus conventional ovarian stimulation for in vitro fertilization: a multicenter, randomized, controlled, assessor-blinded, phase 3 noninferiority trial. Fertil Steril 2017;107:387–396.e4.

10. Qiao J, Zhang Y, Liang X, Ho T, Huang H-Y, Kim S-H, Goethberg M, Mannaerts B, Arce J-C. A randomised controlled trial to clinically validate follitropin delta in its individualised dosing regimen for ovarian stimulation in Asian IVF/ICSI patients. Hum Reprod 2021;36:2452–2462.

11. Robertson I, Chmiel FP, Cheong Y. Streamlining follicular monitoring during controlled ovarian stimulation: a data-driven approach to efficient IVF care in the new era of social distancing. Hum Reprod 2021;36:99–106.

12. Siristatidis CS, Hamilton MP. What should be the maximum FSH dose in IVF/ICSI in poor responders? J Obstet Gynaecol 2007;27:401–405.

13. van Tilborg TC, Torrance HL, Oudshoorn SC, Eijkemans MJC, Koks CAM, Verhoeve HR, Nap AW, Scheffer GJ, Manger AP, Schoot BC, et al. Individualized versus standard FSH dosing in women starting IVF/ICSI: an RCT. Part 1: The predicted poor responder. Hum Reprod 2017;32:2496–2505.

